# Rapid Autopsy Multi-Omic Analysis Identifies Divergent Evolutionary Trajectories and DNA Damage Resistance Mechanisms in FGFR2-Driven Cholangiocarcinoma

**DOI:** 10.64898/2025.11.25.25340882

**Authors:** Ankur Sheel, Anoosha Paruchuri, Julie Reeser, Michele Wing, Raven Vella, Eric Samorodnitsky, Amy Smith, Thuy Dao, Emily Hoskins, Russell Bonneville, Hui-zi Chen, Chunjie Li, Zachary Risch, Patricia Allenby, Aharon Freud, Alexander Fitzthum, Wei Chen, Sameek Roychowdhury

**Affiliations:** Division of Medical Oncology, Department of Internal Medicine, The Ohio State University James Comprehensive Cancer Center, Columbus, OH; Lombardi Comprehensive Cancer Center, Georgetown University, Washington D.C; Division of Hematology and Oncology, Department of Medicine, University of Michigan, Ann Arbor, MI; Division of Hematology and Oncology, Department of Medicine, Medical College of Wisconsin, Milwaukee, WI; Division of Oncology, Washington University School of Medicine, St. Louis, MO; Division of Autopsy Services, Department of Pathology, The Ohio State University James Comprehensive Cancer Center, Columbus, OH

## Abstract

**Background:** Intrahepatic cholangiocarcinoma (iCCA) is an aggressive cancer with poor prognosis. Fibroblast Growth Factor Receptor 2 (FGFR2) alterations, including gene fusions and gain-of-function mutations, occur in ∼10% of iCCA and lead to active FGFR2 signaling. Targeted therapy with an FGFR inhibitor (FGFRi) indeed improves survival, however nearly all patients eventually become resistant to therapy. Moreover, for the broader iCCA population combination of gemcitabine w/ platinum-based chemotherapy remains the standard systemic backbone, yet clinical benefit is limited by acquired resistance. The biological basis of resistance to chemotherapy and the interplay between clonal evolution and chemotherapy resistance remains unknown.

**Methods:** We analyzed 281 whole-exome and 62 transcriptome sequencing samples from 21 patients (11 FGFR2-altered, 10 wild-type) enrolled in a Rapid Autopsy Program. We implemented bioinformatics tools to infer clonal evolution with inferred phylogenetic reconstruction and identified recurrent genomic alterations. We implemented gene set enrichment analyses on tumor transcriptomes from autopsy. DNA damage repair (DDR) mutational signatures were quantified.

**Results:** Tissue collected at the time of autopsy (both liver and metastatic sites) harbored more oncogenic drivers than diagnostic biopsies with recurrent alterations in TP53, BAP1, IDH1/2, KRAS, and PIK3CA. FGFR2-driven tumors displayed fewer truncal oncogenic drivers, but greater clonal dynamics, with recurrent alterations in DDR genes and enrichment of base excision repair (BER) pathways in metastases suggesting evolving DDR vulnerabilities. In one longitudinal case, alternating FGFRi and chemotherapy cycles revealed that resistant FGFR2 clones emerged under targeted therapy, but were subsequently suppressed by platinum rechallenge, highlighting the vulnerability of these subclones.

**Conclusions:** FGFR2-driven iCCA follows an evolutionary trajectory marked by DDR pathway reprogramming. DDR mutational signatures can emerge as biomarkers for therapy resistance. These data highlight opportunities to integrate FGFR inhibition with DDR-targeted agents and supports the idea of platinum rechallenge to overcome resistance.

## INTRODUCTION

Intrahepatic Cholangiocarcinoma (iCCA) is a rare and lethal malignancy with a 5-year overall survival rate of <10% [1]. Until 2022, the chemotherapy backbone for standard of care therapy for iCCA patients was gemcitabine and cisplatin. This regimen conferred an Overall Survival (OS) and Progression Free Survival (PFS) of 11.5 and 5.7 months, respectively [2]. The addition of immunotherapy, durvalumab or pembrolizumab, to chemotherapy improved OS and PFS by a meager 1.3 and 1.5 months, respectively [3, 4]. Despite such a small benefit, the combination of gemcitabine, cisplatin and durvalumab is currently first line therapy for the treatment of iCCA.

The discovery of actionable genomic driver mutations, such as Fibroblast Growth Factor Receptor 2 (FGFR2), in iCCA has transformed clinical management for iCCA such that targeted therapies against FGFR2 are FDA approved in the second line setting. FGFR2 represents a family of receptor tyrosine kinases that control critical physiologic processes, including cell proliferation, survival, growth arrest, differentiation, migration, and apoptosis [5]. Genomic alterations in the FGFR family, including point mutations, gene fusions, alternative splicing, and copy number amplifications are prevalent in ∼10% of iCCA [6]. *FGFR2* aberrations result in dysregulation of these critical physiologic processes, thereby driving iCCA tumorigenesis [5]. In 2020, the FGFR inhibitor (FGFRi) pemigatinib (Pemazyre) was FDA approved for second-line treatment for iCCA patients with *FGFR2* rearrangements as it improved OS from 11.5 months to 21.1 months and progression free survival from 5.7 to 6.9 months compared to standard of care therapy [7].

Despite improved survival outcomes with Pemigatinib, nearly all patients ultimately progress. Previous studies have demonstrated that resistance mechanisms to FGFRi in iCCA include acquired secondary FGFR2 kinase domain mutations [8], bypass signaling via parallel receptor tyrosine kinases including EGFR [9, 10] and EMT programs [11]. Beyond these established pathways, work in lung and ovarian cancer models has further shown that FGFR signaling intersects with DNA damage response (DDR) biology, influencing susceptibility to platinum agents and revealing potential synergy between FGFRi and DNA-damaging chemotherapy [12, 13]. Together, these observations suggest a deeper mechanistic interrogation of how FGFR signaling, DDR pathways, and clonal evolution co-determine therapeutic trajectories in iCCA.

Research autopsy is a powerful tool for studying tumor heterogeneity, evolution, and acquired resistance in multiple cancer types by providing multi-tissue sampling within the same patient. Traditional single-timepoint biopsies capture only a narrow slice of tumor ecology. Rapid research autopsy overcomes this constraint, enabling systematic, multi-site sampling within patients to profile primary tumors and anatomically distant metastases under real-world treatment histories. This design uniquely resolves heterogeneity, temporal evolution, and therapy-conditioned selection pressures. Research derived from rapid autopsy has successfully elucidated novel resistance mechanisms to immunotherapy in melanoma [14], tumorigenesis of gallbladder cancers [15], clonal divergence in pancreatic cancers [16] therapy resistance in small cell lung cancer [17] and mechanisms of FGFRi resistance in cholangiocarcinoma [10].

Here, we leverage a rapid-autopsy cohort of 21 metastatic iCCA patients (11 FGFR2-altered; 10 FGFR2-WT) profiled through multi-region whole-exome sequencing (WES) and transcriptome sequencing. Importantly, all FGFR2-altered patients received FGFR-directed therapy (pemigatinib or infigratinib) in addition to standard chemotherapy, allowing us to study evolutionary dynamics across treatments. Of note, the patients in this cohort received therapy prior to the approval of the addition of durvalumab in the first line setting. We present a comprehensive genomic and transcriptomic analysis using samples from tissue biopsy and rapid autopsy from both FGFR altered and WT iCCA patients. Our study is one of the first to investigate differences in clonality and transcriptional programs across primary and metastatic sites in FGFR wild type and FGFR altered tumors from patients who were treated with both chemotherapy and FGFRi. Our analysis additionally reveals key differences between hepatic lesions and distant metastases offering insights into the molecular heterogeneity and adaptive mechanisms of iCCA. This integrative framework allows us to bridge clinical observations with genomic insights thereby advancing the application of precision oncology to patient care.

## METHODS

### Rapid Research Autopsy Cohort

Informed consents were obtained from 21 patients with metastatic iCCA to participate in an institutional review board–approved clinical study for tumor profiling by next-generation sequencing and body donation (NCT02090530). All samples were assigned internal study-specific identifiers that were fully de-identified and not linked to any patient-identifying information; these sample IDs were not known to anyone outside the research team. Complete methodology in handling specimens is as previously described [17].

### Sample preparation for sequencing

Samples with tumor cell content greater than 30% and without substantial necrosis were selected for next-generation sequencing analyses. Genomic DNA and total RNA were extracted using Qiagen kits per manufacturer’s protocol. Complete library preparation and reagents utilized as previously described [17].

### Whole Exome Somatic mutation SNV and CNV analysis

These bioinformatics analyses were performed as previously described [17]. Briefly, sequencing reads were aligned to human genome build 19 (hg19) using Burrows-Wheeler Aligner (bwa)[18], deduplication with Picard (https://github.com/broadinstitute/picard), and base quality score recombination and realignment around insertion and deletion (indels) with GATK HaplotypeCaller [19]. Variants were called with VarScan2 [20], Mutect2 [19] and allele-specific copy number variations (CNVs) with VarScan2 [20], CONTRA [21], and FishingCNV [22].

### Circulating Tumor DNA Sequencing

Circulating tumor DNA (ctDNA) sequencing was isolated using QIAamp Circulating Nucleic Acid Kit (Qiagen) per manufacturer’s protocol as previously described [17].

### Driver Mutation and Clonality Analysis

Driver mutations were annotated using OncoKB [23]. Clonal interference was performed using Canopy [24] and mutational signatures were inferred with ASCETIC [25]. Bradley-Terry modeling was used to estimate relative ordering of mutations in the phylogeny branches of autopsy patients with iCCA as previously described [17]. COSMIC was utilized to identify signatures associated with SNVs [26].

### Transcriptome Analysis

These analyses were performed as previously described [17]. Briefly, raw FASTQs were aligned to hg19 using STAR-2.7.11b[27]. Resulting BAMs were sorted using samtools-1.21[28]. Fusions were called using Arriba v2.4.0[29] and STAR-Fusion v1.13.0[30]. Gene expression was calculated using Salmon-1.10.1 [31] and CuffLinks-2.2.1[32]. Differentially expressed genes were identified using DESeq2 [33]. DDR pathway genes were derived from GSEA [34]. Expression of DDR genes was compared using TPMs.

### Statistical Analysis

All statistical analyses were conducted in R 4.4.0.

### Supercomputing

All analyses were done on the Ohio Supercomputer Center’s (https://www.osc.edu/) Pitzer and Ascend clusters.

## Data Availability

Whole exome and transcriptome samples used in this study are available upon reasonable request.

## RESULTS

### Patient Cohort and Sample Collection

Demographic and clinical information of the 21 patients with iCCA who underwent research autopsy are presented in Figure 1 and Supplementary Figure 1. In total, we performed whole-exome sequencing (WES) on 281 samples and RNA-seq on 62 samples including archival liver biopsy specimens as well as autopsy specimens from the liver, lymph nodes, and distant metastatic sites (Supplementary Figure 2). Lesions spanned intrahepatic and extrahepatic locations including lung, breast, pericardium, omentum, bone, spleen, kidney and ovaries (Figure 1B). All patients had stage IV disease at the time of diagnosis and received multiple lines of treatments, including first-line gemcitabine with cisplatin as autopsies were performed before the approval of durvalumab. 11 patients in our cohort had FGFR2 alterations and 10 patients in our cohort were FGFR-WT. It is important to note in this study that all 11 patients with FGFR2 alterations received FGFR directed therapy with either Pemigatinib or Infigratinib (Figure 1C). Complete treatment histories and sampling timelines were documented for each patient (Figure 1C). Pretreatment samples were included for WES and RNA-seq when available.

**Figure 1.**
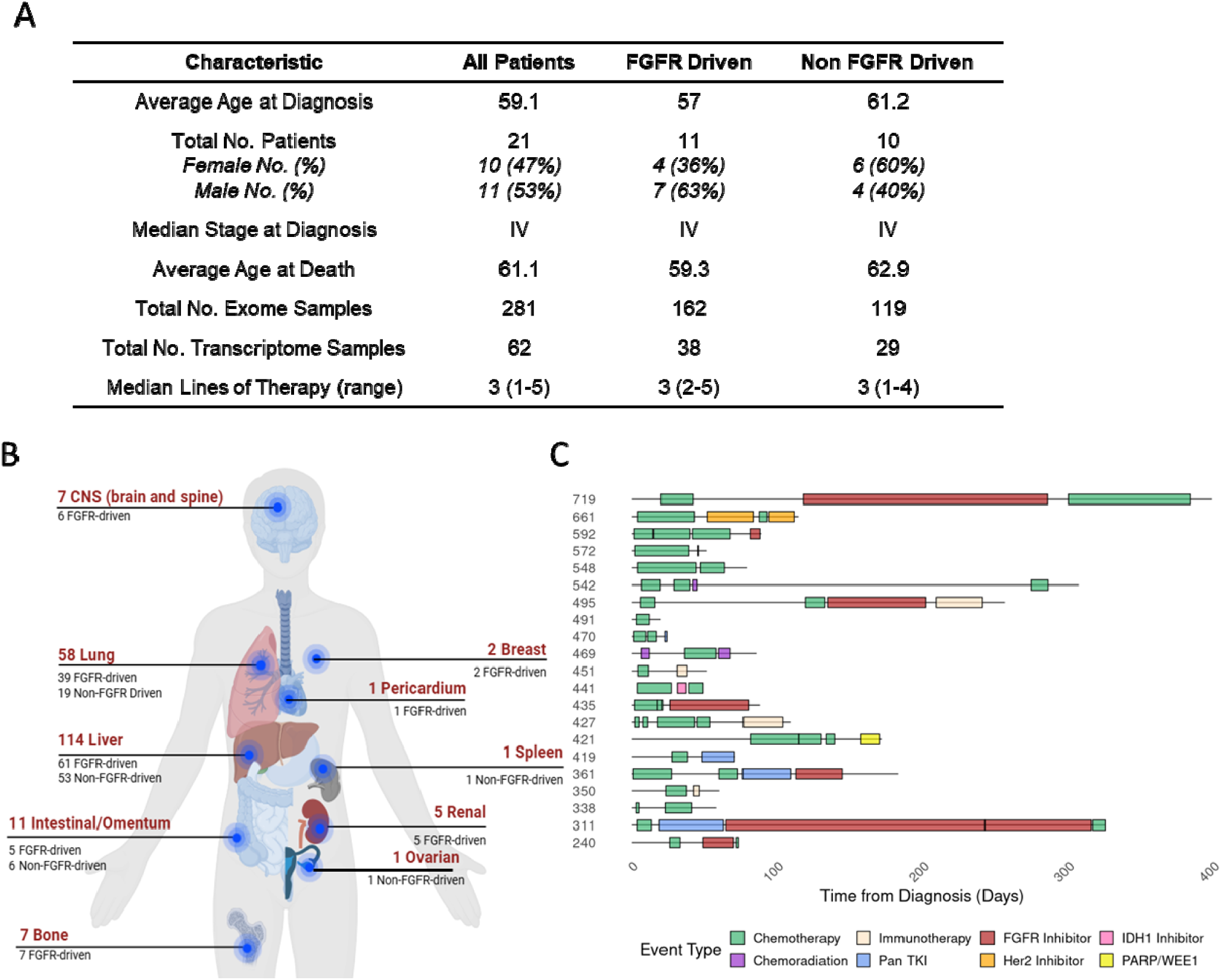
iCCA Cohort Sample Descriptions and Patient Characteristics. (A) Baseline characteristics of patients with FGFR driven and non FGFR driven iCCA. (B) Anatomical distribution of lesions collected from rapid research autopsy with stratification by FGFR-driven versus non FGFR-driven cases. (C) Timeline of treatment exposure by patient. Each bar represents a patient’s therapeutic timeline from diagnosis to death. Length of boxes indicate duration of treatment and gaps between boxes represent time off therapy.

### Genetic Alterations Across Primary and Metastatic Lesions

To investigate anatomical and temporal tumor evolution, we performed multi-site whole-exome sequencing across liver biopsies, lymph nodes, and distant metastases from patients with cholangiocarcinoma (Figure 2). The mean number of somatic mutations, including insertion-deletions (indels) was 571 per tumor, the tumor mutational burden ranged from 6.1 to 19.4 mutations per megabase (Mb) (Figure 2A and Supplemental Figure 3A). To determine the contribution of somatic mutations to tumorigenesis, we annotated SNVs based on oncogenicity as determined by OncoKB [23] and limited our subsequent analysis to SNVs and indels classified as “Oncogenic” and “Likely Oncogenic.” We determined that samples collected at the time of autopsy including liver and metastatic lesions carried significantly more driver mutations than archival liver biopsies (****p < 0.0001; **p < 0.01) (Figure 2B), which is consistent with clonal diversification during disease progression. Oncoplot analysis confirmed recurrent SNVs in TP53, BAP1, IDH1/2, KRAS, and PIK3CA, with FGFR2 fusions confined to the FGFR-driven group (Figure 2C). Notably, FGFR driven tumors were found to have fewer oncogenic drivers compared to non-FGFR driven tumors (Supplementary Figure 3B), suggesting a distinct mechanism for genomic complexity and disease progression. Comparing our driver list with TCGA and other published iCCA cohorts [35, 36], we identified mutations in 29 novel genes that have not previously been reported as drivers of iCCA in prior studies (Supplemental Figure 3C). Interestingly, the novel genes identified in our study contained genes, such as BLM, CHEK1, CHEK2, FANCL, PTCH1, RAD50, RECQL, and XRCC2, which are all implicated in DNA damage repair and mitotic stability [34]. Although most driver mutations were conserved across all sampled sites within individual patients (Figure 2D), a number of private mutations were uniquely identified in liver and metastatic lesions at autopsy, indicating regional clonal diversification. Notably, we identified a core set of 21 shared driver SNVs present across all sample types in each patient (Figure 2E and Supplementary Figure 3D), suggesting these mutations may underpin essential oncogenic programs and reflect potential points of tumor dependency or "oncogene addiction." Together, these findings highlight the importance of multi-region sampling and integrative mutational analysis to capture a dimensional spectrum of tumor heterogeneity in metastatic CCAs.

**Figure 2.**
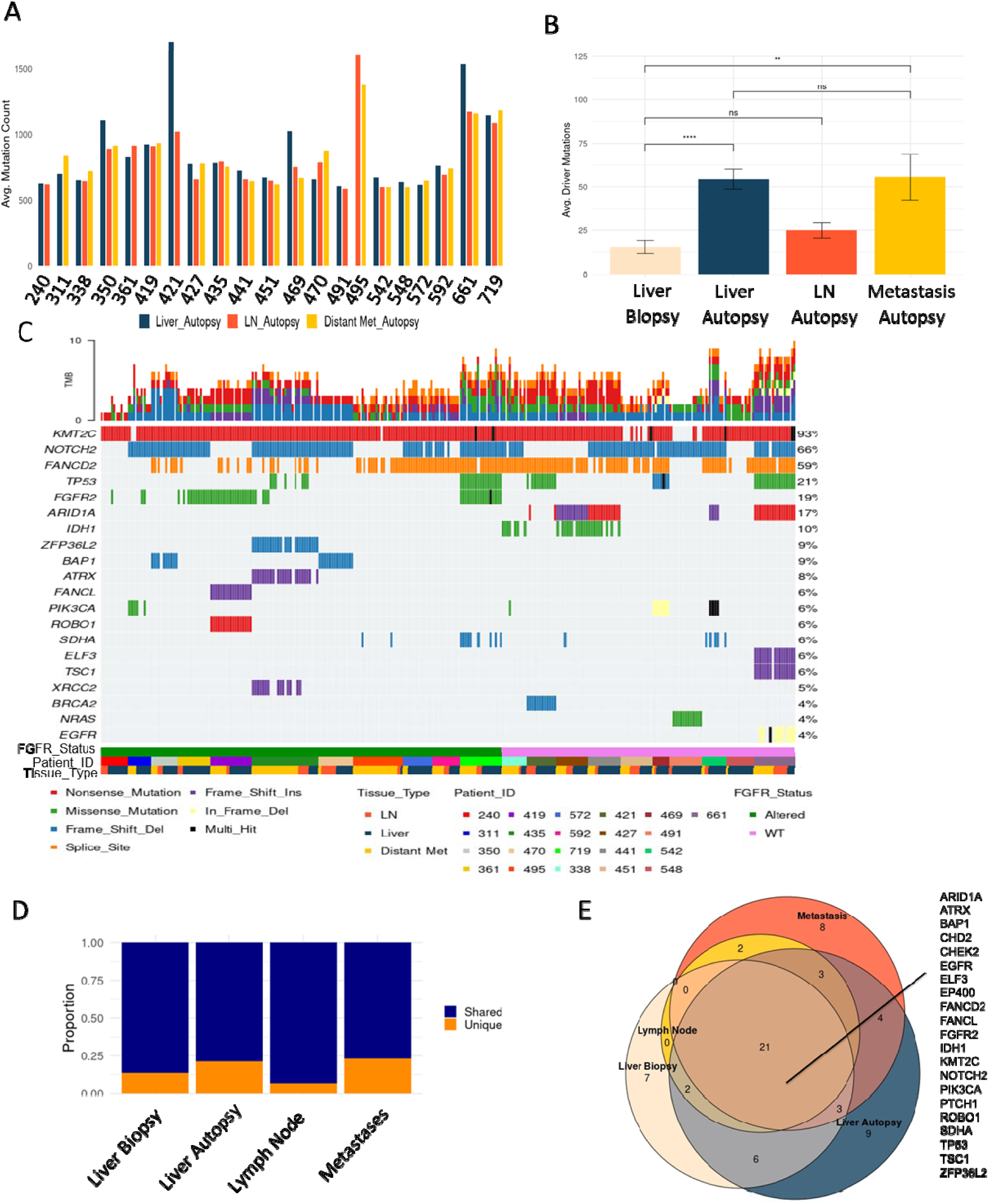
Genomic landscape and mutational burden across primary and metastatic lesions. (A) Bar plot showing average mutation count per sample across liver, lymph node and distant metastases obtained at the time of autopsy for each patient. (B) Comparison of average driver mutation burden by sample type. Liver Autopsy and Metastases samples had significantly greater average number of driver mutations compared to Liver Biopsy samples (****p < 0.0001; **p < 0.01; ns = not significant). (C) Oncoplot summarizing key oncogenic mutations across patients. Additional annotations below include treatment type, tissue type and time of sampling (biopsy vs autopsy). (D) Proportion of mutations classified as shared and unique across tissue types. Share mutations are defined as those detected in ≥2 tumor sites within a patient whereas unique mutations are exclusive to a single sample type. (E) Venn diagram showing overlapping oncogenic drivers across sample types within the autopsy cohort. A core set of 21 mutations were shared across all anatomical sites within a single patient.

### Divergent Copy Number Alterations Reflect Site-Specific Clonal Evolution

We next evaluated genome-wide somatic copy number alterations (CNAs) across tumor sites to assess the extent and divergence of chromosomal instability in primary and metastatic lesions. Copy number profiles from representative samples demonstrated widespread aneuploidy across all tissue types, with recurrent chromosomal gains and deletions observed across multiple chromosomes in liver biopsy (Supplementary Figure 4A), liver autopsy (Supplementary Figure 4B), lymph node samples from autopsy (Supplementary Figure 4C), and distant metastases from autopsy (Supplementary Figure 4D). When aggregated across all samples, the relative distribution of copy number states (amplified, diploid, and loss) did not significantly differ between tissue types (Supplementary Figure 4E, χ² p = 0.5856), indicating that the global burden of CNAs is comparable across sites. However, focusing specifically on key oncogenic drivers revealed divergent patterns of alterations, copy number alterations in amplified, diploid, or loss states were heterogeneously distributed across primary and metastatic lesions (Supplementary Figure 4F).

Notably, when examining the FANCD2 locus, a gene involved in DNA repair and replication stress response, we observed significant variation in copy number status across tissue types. Liver biopsy samples were predominantly diploid for FANCD2, whereas liver autopsy, lymph node, and metastatic samples showed increased frequencies of both loss and amplification events (Supplementary Figure 4G). This shift in FANCD2 copy number was statistically significant (χ² p = 0.0056), suggesting a role for dynamic modulation of this locus during disease progression and clonal evolution. These findings support the need for multi-site genomic profiling to fully capture the landscape of actionable alterations.

### Clonal Evolution Reveals Distinct Patterns of Intra-Tumoral Heterogeneity

To investigate the clonal diversity of iCCAs, we quantified intra-tumoral heterogeneity using the Shannon Index, a commonly used measure of diversity [37], across distinct sites of disease. We observed a significant increase in clonal diversity in metastases compared to primary liver sites. This suggests that the occurrence of new clones (clonal expansion) co-occurs with metastasis (Figure 3A). Stratifying by FGFR2 fusion status, FGFR2 altered tumors exhibited significantly lower heterogeneity at both autopsy and metastatic sites compared to their FGFR2-wild-type (WT) counterparts (Figure 3B) suggesting that FGFR fusions are truncal in nature. This pattern suggests FGFR2 fusion events dominate the clonal architecture and potentially constrain further clone diversification.

**Figure 3.**
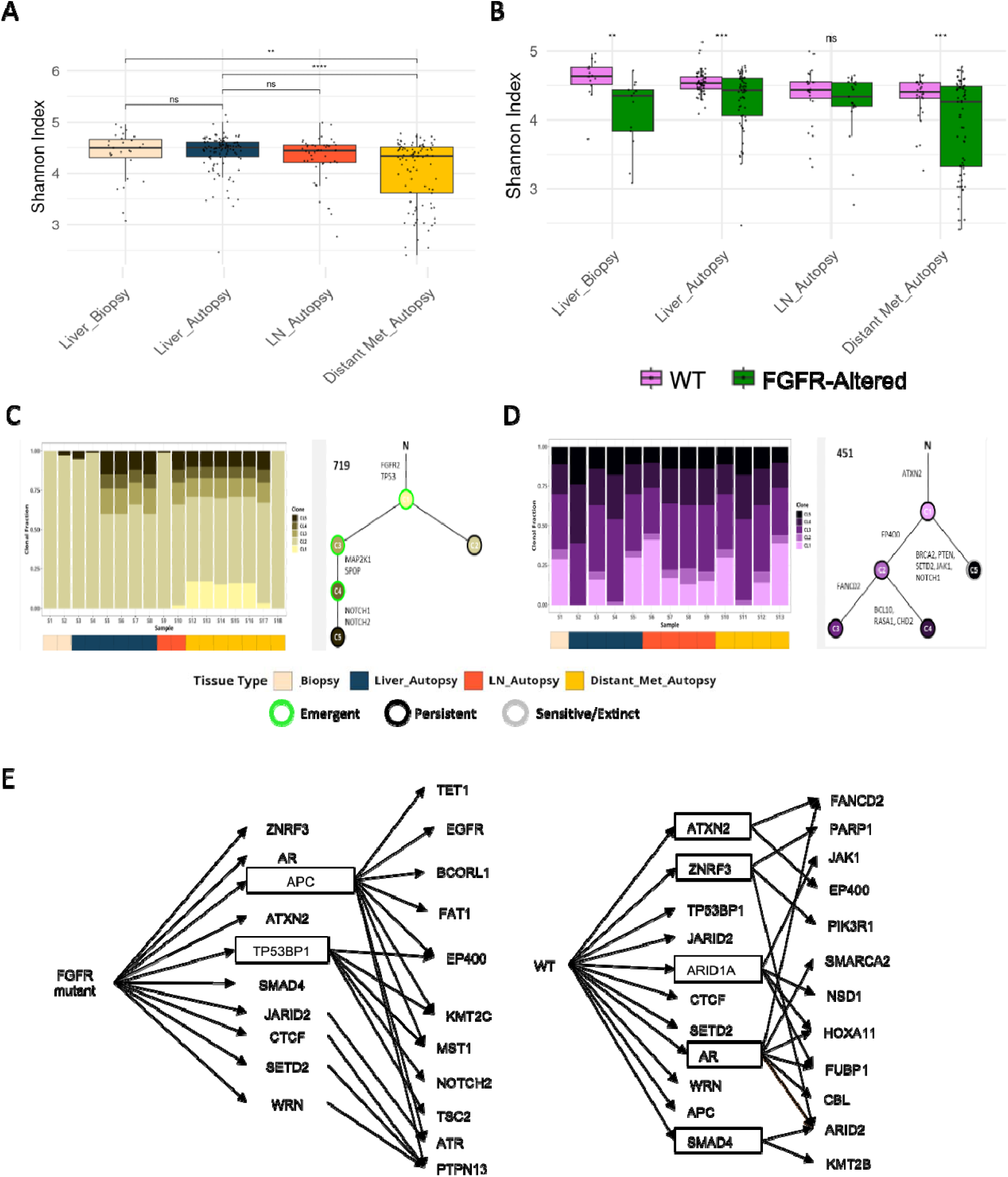
Clonal evolution and DNA damage repair enrichment in FGFR2-driven iCCAs. (A) Shannon index quantifying intra-tumoral heterogeneity across liver biopsy, liver autopsy, and distant autopsy samples across all samples. (B) Shannon index quantifying intra-tumoral heterogeneity in FGFR fusion driven vs FGFR WT liver biopsy, liver autopsy, and distant autopsy samples. Clonal dynamics across tumor sites with trees depicting emergence and extinction of subclones across sites. Bar plots represent clonal prevalence across tissue types (biopsy, liver autopsy, lymph node autopsy and distant metastasis autopsy) for an (C) FGFR driven patient and (D) FGFR WT patient. (E) Phylogenetic trajectory of driver mutations in FGFR2 fusion-positive (green) and FGFR2-wild-type (purple) tumors. Mutations in DNA damage repair genes (e.g., *APC and TP53BP1*) are enriched in FGFR2 fusion cases and emerge along branches, suggesting DNA repair reprogramming during clonal evolution. Mutations in *ATXN2*, *ZNRF3*, *ARID1A*, *AR*, and *SMAD4* are enriched in FGFR WT genes suggesting chromatin remodeling, hormone signaling and TGF-β pathway regulation.

Clonal deconvolution of patient-matched samples further revealed distinct patterns of subclonal emergence and extinction. In an FGFR2 fusion–positive patient, we observed extensive clonal reshaping between biopsy, liver autopsy, lymph node, and distant metastatic samples (Figure 3C), with several emergent clones detected only at later stages. In contrast, an FGFR WT patient demonstrated greater clonal stability with fewer emergent clones (Figure 3D).

In the FGFR2 mutant patient (Figure 3C), a gradual reduction in the number of minor subclones was observed from biopsy to autopsy samples, indicating clonal convergence. Evolutionary tree reconstruction revealed a central ancestral clone harboring FGFR2 and TP53 alterations, with subsequent branching driven by mutations in MAP2K1, SPP2, and NOTCH family genes. In contrast, the WT case (Figure 3D) displayed a more even clonal distribution across samples, suggesting the persistence of multiple subclones through disease progression.

Phylogeny reconstruction and clonal deconvolution of patient-matched samples further revealed distinct patterns of subclonal emergence and extinction. In an FGFR2 fusion–positive patient, we observed extensive clonal reshaping between biopsy, liver autopsy, lymph node, and distant metastatic samples (Figure 3C), with several clones detected only at later stages. In contrast, an FGFR WT patient demonstrated greater clonal stability with fewer emergent clones (Figure 3D). These findings suggest that FGFR2-driven tumors undergo more dynamic clonal evolution compared to FGFR2-WT tumors.

To determine the evolutionary programs favored by FGFR2 fusion-positive vs FGFR WT iCCA, we employed phylogenetic mapping of driver mutations using ASCETIC [25]. This analysis revealed a divergent spectrum of evolutionary pressures between FGFR2 fusion–positive and WT tumors (Figure 3E, Supplementary Figure 5 and Supplementary Figure 6). In FGFR2 fusion–positive tumors, DNA damage repair genes—including *APC* and *TP53BP1*—frequently appeared along late-branching subclones, consistent with ongoing reprogramming of DDR pathways during tumor progression. Conversely, FGFR WT tumors exhibited enrichment of mutations in *ARID1A*, *SMAD4*, *AR*, and *ZNRF3*, implicating chromatin remodeling, hormone receptor signaling, and TGF-β pathway alterations as dominant evolutionary drivers. These results indicate clonal evolution in FGFR2 fusion positive patients favors DNA repair–associated alterations, potentially contributing to therapy resistance and metastatic competence.

### DNA damage pathway dysregulation in metastases

Given our previous observations, we performed mutational signature analysis across spatially distinct tumor lesions. Analysis of COSMIC single base substitution (SBS) signatures revealed marked differences in mutational processes by site of disease (Figure 4A–B). Distant metastases exhibited a higher proportion of SNV signatures associated with mismatch repair (MMR) deficiency, defective homologous recombination, and platinum chemotherapy exposure, suggesting defective DDR in these lesions. These results are consistent with our results regarding preferential evolution towards deficient DNA damage repair in FGFR2 fusion positive patients.

**Figure 4.**
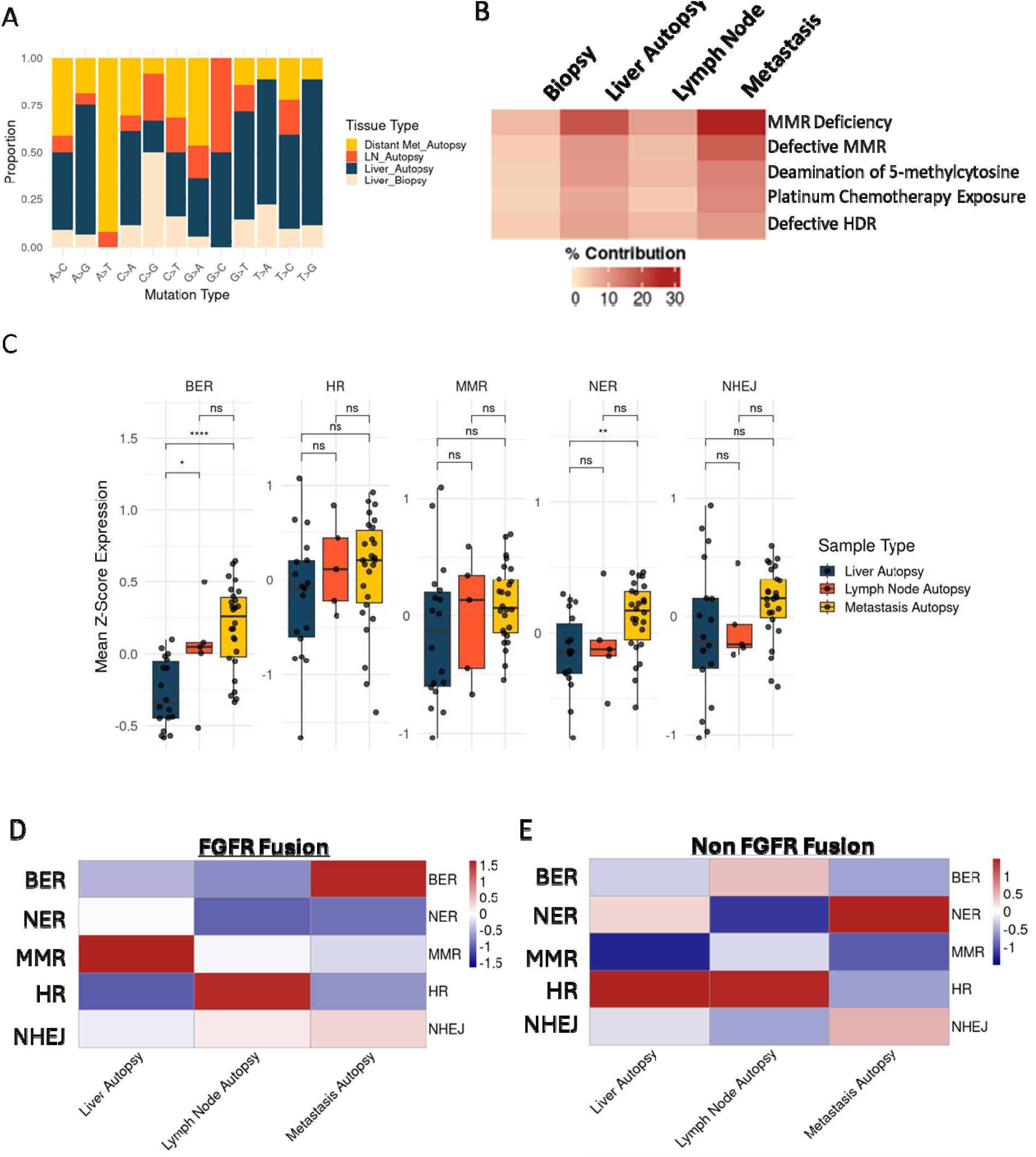
Defects in base excision repair (BER) drive DNA damage accumulation in metastatic lesions. (A) Proportion of mutational signatures across all samples, stratified by tissue type (liver biopsy, liver autopsy, lymph node autopsy, and distant metastasis autopsy). (B) Heatmap of COSMIC single base substitution (SBS) signatures across tumor samples, highlighting elevated SNV signatures associated with MMR deficiency and defective HDR in distant metastatic samples. (C) Mean z-scored expression of genes in five DNA repair pathways—base excision repair (BER), homologous recombination (HR), mismatch repair (MMR), nucleotide excision repair (NER), and non-homologous end joining (NHEJ)—across liver autopsy, lymph node autopsy, and metastasis autopsy samples. (****p < 0.0001, Kruskal–Wallis test). (D) Heatmap summarizing DNA repair pathway expression across sample types in FGFR Fusion and (E) FGFR-WT patients.

Next, we performed pathway-level transcriptomic profiling to confirm our results from genomic alteration analysis, clonal evolution analysis, and mutational signatures indicating deficient DNA damage repair is a hallmark of metastasis. Transcriptome analysis of DNA damage response (DDR) pathways demonstrated significant upregulation of base excision repair (BER) in metastases compared to both liver autopsy and lymph node lesions (Figure 4C, Supplementary Figure 7). No significant differences were observed in MMR, nucleotide excision repair (NER), or non-homologous end joining (NHEJ) across sites. When stratified by FGFR2 fusion status, this upregulation in BER related gene expression in metastatic lesions was most prominent in FGFR2 fusion-positive samples (Figure 4D), while non-fusion samples exhibited enrichment in NER (Figure 4E). These findings suggest that enhanced BER may represent compensatory mechanisms in response to accumulated DNA damage during metastatic spread, particularly in the context of FGFR2-driven disease.

### Chemotherapy suppresses genomically unstable clones after FGFR resistance

Longitudinal genomic and radiographic analysis of an FGFR2 fusion–positive iCCA patient revealed marked clonal evolution in response to cyclical therapy. Initial gemcitabine/cisplatin chemotherapy produced radiographic regression, followed by ∼14 months of disease control on FGFR inhibition with Infigratinib (Figure 5A). Over time, however, resistant subclones harboring acquired FGFR2 kinase domain mutations (V565I, N550K, N550T) emerged, accompanied by rising FGFR2 fusion variant allele frequency and clinical progression (Figure 5B). Notably, reintroduction of chemotherapy not only led to a radiographic response but also was associated with a fall in both the dominant FGFR2 fusion clone and the associated resistant subclones. These findings align with our broader clonality analysis showing defective DDR and suggest that chemotherapy retains efficacy against these unstable and heterogeneous clones.

**Figure 5.**
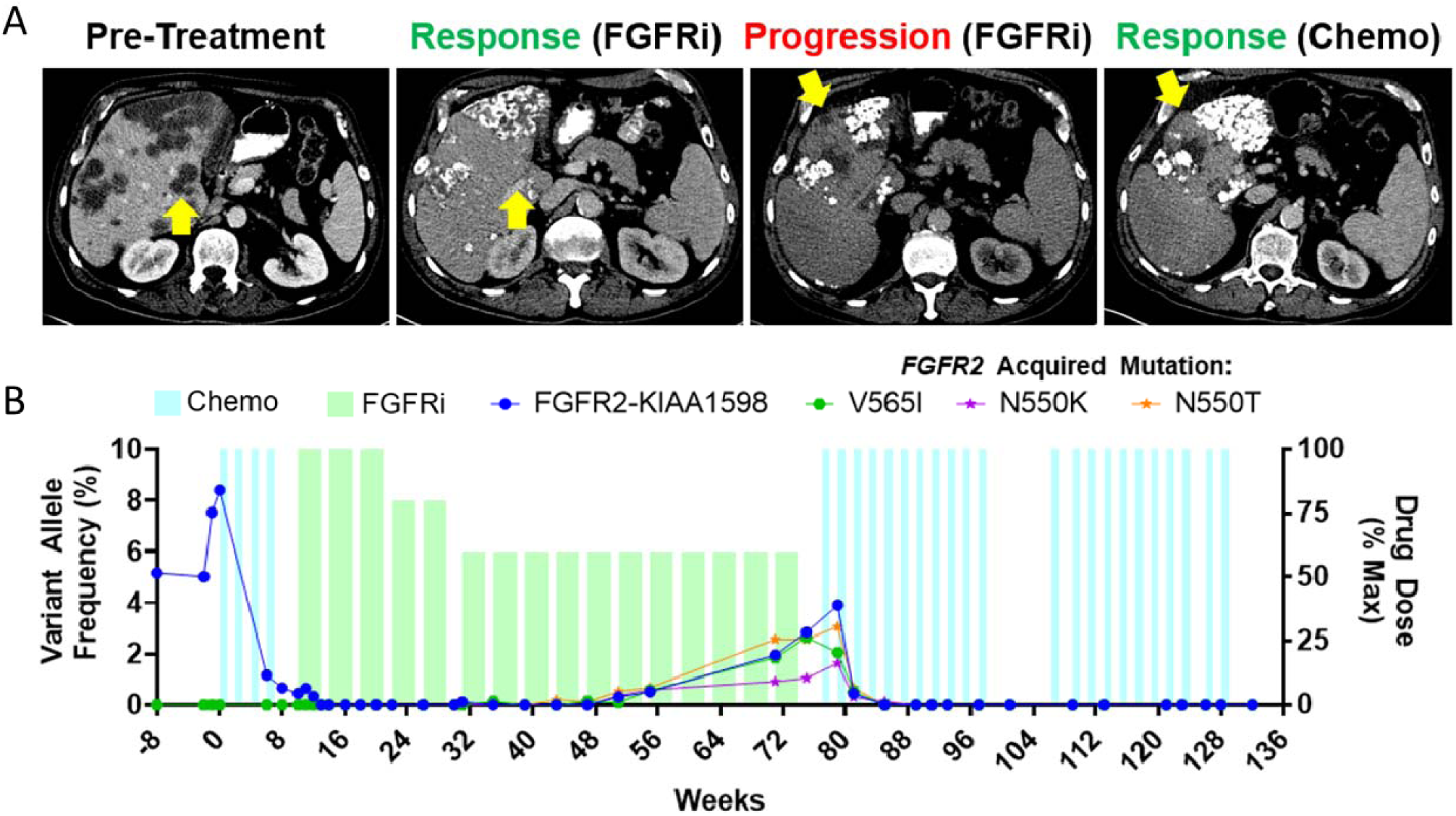
Serial FGFR cfDNA monitoring demonstrates potential benefit of cyclical therapy in cholangiocarcinoma. A) *FGFR2*-fusion positive cholangiocarcinoma patient received 2 months of gemcitabine/cisplatin chemotherapy, changed therapies to FGFRi (infigratinib) until radiographic disease progression after ∼14 months. At this time, disease was successfully treated with second exposure to gemcitabine and cisplatin. B) Serial monitoring of *FGFR2* fusions and point mutations showed a decrease in *FGFR2-KIAA1598* VAF with initial treatment, and an increase in VAF which preceded radiographic disease progression. Resistant subclones of FGFR occurred while on infigratinib, and these were eliminated with re-introduction of gemcitabine and cisplatin.

## DISCUSSION

In this comprehensive autopsy-based study of iCCA, we leveraged whole-exome and transcriptome sequencing across a unique set of spatially and temporally distinct tumor samples from 21 patients to dissect the molecular evolution patterns and associate these findings with therapy resistance in FGFR2 fusion–driven and wild-type tumors. Our data provides new insights into the complex mechanisms driving disease progression and therapy resistance and suggests an evolutionary model where therapeutic pressure against FGFR2 reshapes DNA repair pathways. Furthermore, our findings highlight the potential of chemotherapy to target unstable resistant clones following FGFR inhibitor failure and suggest that DDR pathway activity may serve as therapeutic targets or biomarkers of resistant therapy.

Our multi-site genomic analysis revealed a notable expansion in the landscape of driver mutations in cholangiocarcinoma. By comparing against established cohorts (TCGA, Boerner et al., Zhou et al.), we identified 29 novel driver genes not previously implicated in iCCA, substantially broadening the catalog of putative oncogenic drivers. Interestingly, many of these novel alterations occurred in genes associated with DNA damage repair and chromosomal stability—including *BLM*, *CHEK1*, *CHEK2*, *FANCL*, *RAD50*, *PTCH1*, *RECQL*, and *XRCC2*—highlighting an underappreciated role for genomic maintenance mechanisms in driving iCCA. These findings suggest that defective DDR may be a central axis of tumor progression, particularly in the metastatic setting.

Strikingly, despite regional diversification and the emergence of private mutations in liver and distant metastatic lesions, we observed a core set of 21 driver SNVs that were consistently shared across all sampled tumor sites within individual patients suggesting that these alterations may constitute essential oncogenic programs that remain active throughout disease progression. Their conservation across space and time supports the hypothesis of tumor “addiction” to these drivers and underscores their potential as therapeutic targets. This hypothesis warrants further experimental validation.

While no computational tool exists that incorporates the presence of fusions in clonality determination, interpretation of the Shannon index from samples demonstrates that FGFR2 samples have decreased clonal diversity compared to FGFR WT tumors suggesting that the fusion is truncal. Consistent with this, FGFR2 fusion–positive tumors exhibited fewer total oncogenic drivers than FGFR WT cases, implying that the fusion acts as an early, dominant oncogenic event that reduces selective pressure for additional founder mutations. These observations are in line with previous observations demonstrating that FGFR mutations and fusions occur early in tumorigenesis [38]. Despite the lower baseline heterogeneity, FGFR2-driven tumors exhibited dynamic clonal evolution under therapeutic pressure, with the emergence of new subclones enriched in DDR alterations—particularly in genes such as APC and TP53BP1. These findings suggest a unique evolutionary trajectory for FGFR-driven tumors resulting in increasingly defective DDR over time, especially following selective pressure from FGFR inhibition. Inversely, in FGFR-WT tumors, that are treated with chemotherapy, the evolutionary trajectory results in mutations in chromatin remodelers suggesting perhaps an epigenetic mechanism of resistance.

The enrichment of somatic mutations in genes critical to DDR—such as *BLM*, *CHEK1*, *CHEK2*, *FANCL*, *RAD50*, *RECQL*, and *XRCC2*—suggest that genomic instability is a fundamental feature of metastatic iCCA. These genes play essential roles in DDR repair and replication fork protection, and their disruption can compromise genome integrity, promote chromosomal aberrations, and fuel subclonal diversification. Our multi-site sequencing data revealed a higher burden of known driver mutations [39] in autopsy samples compared to diagnostic biopsies, consistent with ongoing clonal evolution. Moreover, phylogenetic reconstruction and elevated Shannon indices in both FGFR-driven and wild-type tumors further support an active process of genomic diversification during disease progression. Together, these findings introduce a model where loss of genome maintenance mechanisms drives the accumulation of oncogenic alterations and enables the emergence of aggressive subclones that underpin therapeutic resistance in metastatic sites.

Our multi-region sequencing revealed a high burden of copy number alterations (CNAs) across all disease sites. Notably, the FANCD2 locus, central to the Fanconi anemia DNA repair pathway, exhibited marked variation in copy number across sample types, with amplification or loss observed more frequently in autopsy and metastatic lesions compared to liver biopsies. These changes correlated with transcriptional upregulation of base excision repair (BER) genes and increased representation of DDR mutational signatures in FGFR2 fusion–positive metastases. Collectively, these findings highlight the role of DNA repair reprogramming in metastatic evolution and suggest that metastatic clones experience and potentially exploit DNA damage for adaptive advantage.

Resistance to FGFR-targeted therapy in cholangiocarcinoma is frequently mediated by the emergence of gatekeeper or kinase domain mutations in the FGFR2 gene, which diminish drug binding and confer therapeutic escape. Prior studies have documented polyclonal resistance via parallel evolution of distinct FGFR2 mutations-including V564F/I, N550K, and K660M-within individual patients, highlighting the genomic plasticity of FGFR-driven tumors under selective pressure[10]. In our study, we observed a similar phenomenon in a longitudinal case of an FGFR2 fusion–positive patient treated with infigratinib, where sequential cfDNA profiling revealed the emergence of FGFR2-KIAA1598 point mutations coinciding with clinical and radiographic disease progression. The reintroduction of platinum-based chemotherapy led to a marked reduction in variant allele frequencies of these resistant subclones, suggesting that chemotherapy can resensitize tumors by suppressing or eliminating genomically unstable lineages. This finding supports the broader trend observed in our clonality analysis, where DDR mutated metastatic clones, particularly in FGFR2 fusion–positive tumors, harbored DDR pathway alterations and were likely vulnerable to DNA-damaging agents. Overall, these findings reinforce known mechanisms of FGFR resistance while also providing novel evidence that chemotherapeutic re-challenge may serve as an effective salvage strategy following FGFR inhibitor failure.

## Limitations

While this study provides unprecedented spatial and temporal resolution of iCCA evolution, several limitations must be acknowledged. First, our sample size, particularly for transcriptomic comparisons, was limited by RNA quality and availability in autopsy tissues. Second, clinical heterogeneity—including differences in prior therapies, timing of biopsies, and treatment durations—introduces variability that may confound some comparisons. Finally, our analyses rely on bulk WES/RNA and therefore cannot resolve cell-to-cell heterogeneity, the co-occurrence of subclonal CNAs within individual cells. Integrating single-cell whole-genome sequencing methods would more accurately refine phylogenies, and test whether DDR-altered, FGFRi-resistant lineages are present at low frequency prior to therapy or arise as a result of therapy.

## Conclusions

This study establishes a multi-site, temporal framework for studying tumor evolution through rapid autopsy, demonstrating how longitudinal sampling across sites of disease and therapy exposure can reveal dynamic evolutionary programs that are not evident in single-timepoint analyses. We demonstrate that FGFR2-driven iCCA follows a distinct evolutionary path marked by truncal oncogenic events and adaptive mutations in DDR. Our study introduces an evolutionary model of mutations in DDR genes in FGFR2-driven cholangiocarcinoma, showing that therapeutic pressure against FGFR2 reshapes DNA repair pathways. Furthermore, our findings highlight the potential of chemotherapy to target unstable resistant clones following FGFR inhibitor failure and suggest that DDR pathway activity may serve as therapeutic targets or biomarkers of resistant therapy. Beyond its biological findings, this work highlights the clinical potential of integrating post-mortem genomics with real-world treatment histories to guide precision oncology. Together, these data advocate for a more personalized, evolution-informed approach to the treatment of advanced cholangiocarcinoma, integrating longitudinal molecular profiling with adaptive therapeutic sequencing.

## Data Availability

All data produced in the present study are available upon reasonable request to the authors

## Acknowledgements

This work was supported by Institutional funds from The Ohio State University Institutional Funds, Pelotonia Institute of Immuno-Oncology and philanthropy. We thank the Ohio Supercomputer Center (https://www.osc.edu/) for providing the computational space on which this work was performed.

## Notes

### Competing Interest Statement

The authors have declared no competing interest.

### Funding Statement

This study was funded by Ohio State institutional funds, Pelotonia Institute of Immunooncology and philanthropy funds.

### Author Declarations

Ethics committee/IRB of The Ohio State University Wexner Medical Center gave ethical approval for this work

