## Supplementary Figures for "Rapid Autopsy Multi-Omic Analysis Identifies Divergent Evolutionary Trajectories and DNA Damage Resistance Mechanisms in FGFR2-Driven Cholangiocarcinoma"

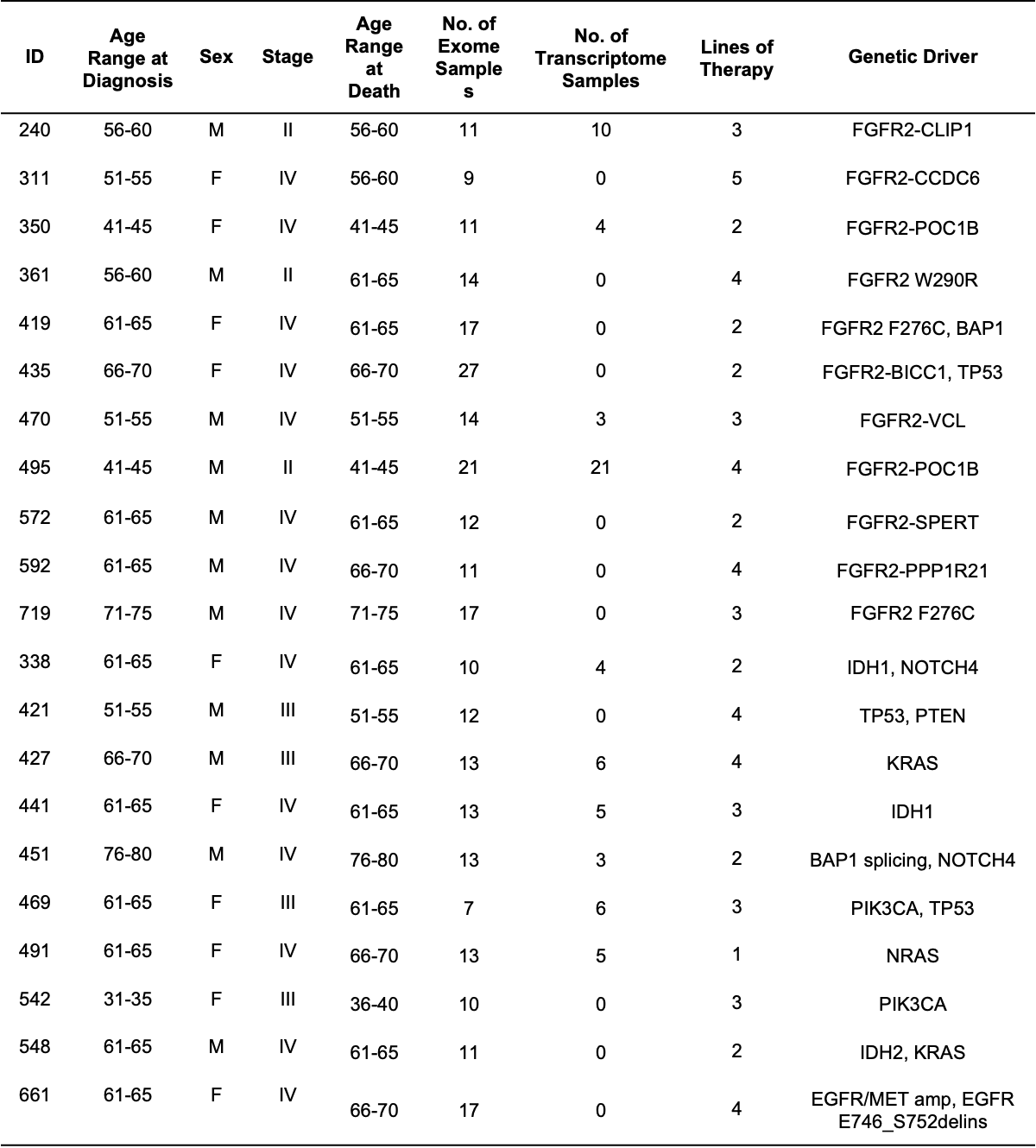

**Supplementary Figure 1.** Autopsy Cohort Characteristics by Patient ID

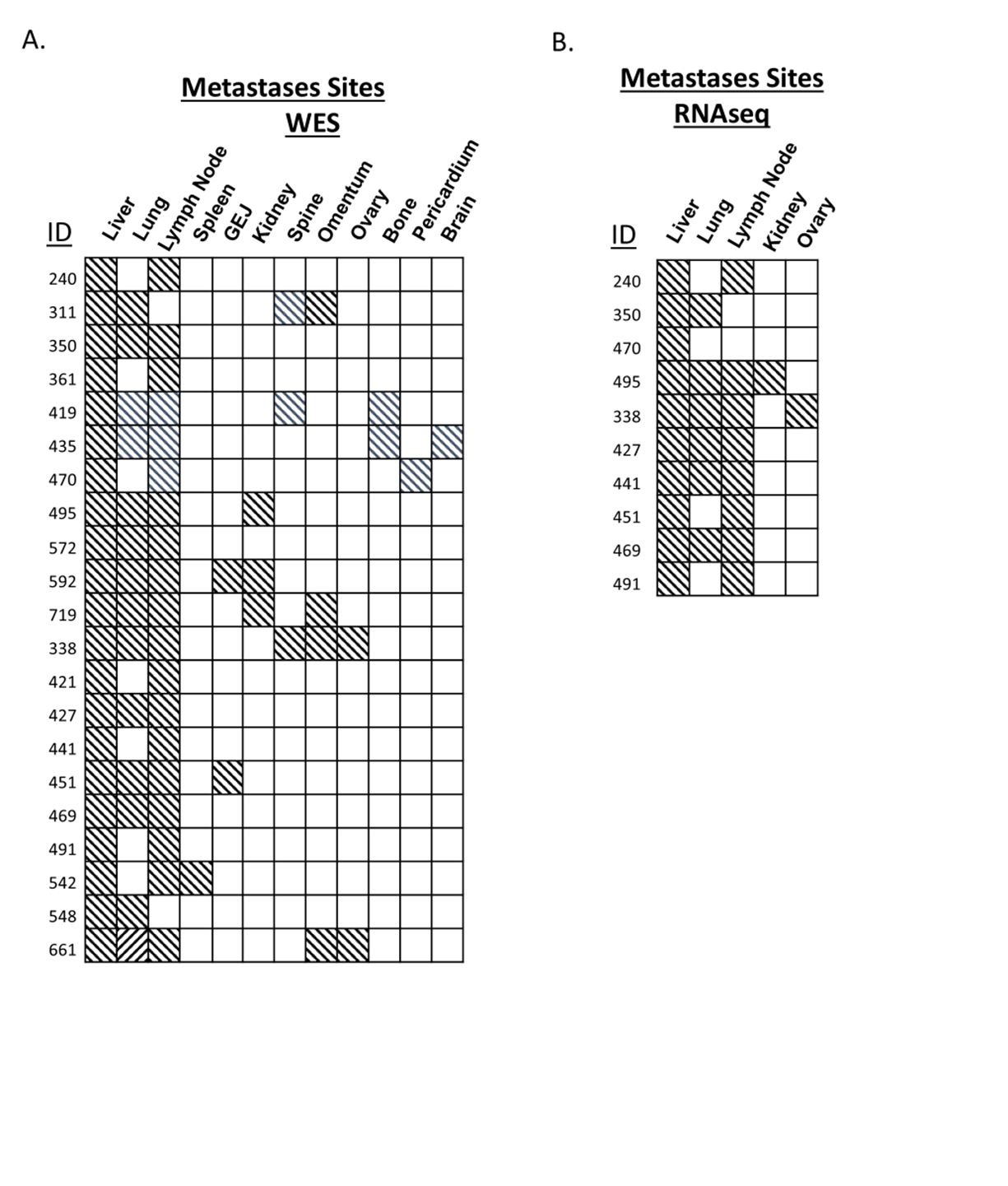

**Supplementary Figure 2.** Distribution of metastatic sites by Patient ID sampled for multi-omic profiling in rapid autopsy cohort for Whole Exome Sequencing (A) and RNA-sequencing (B)

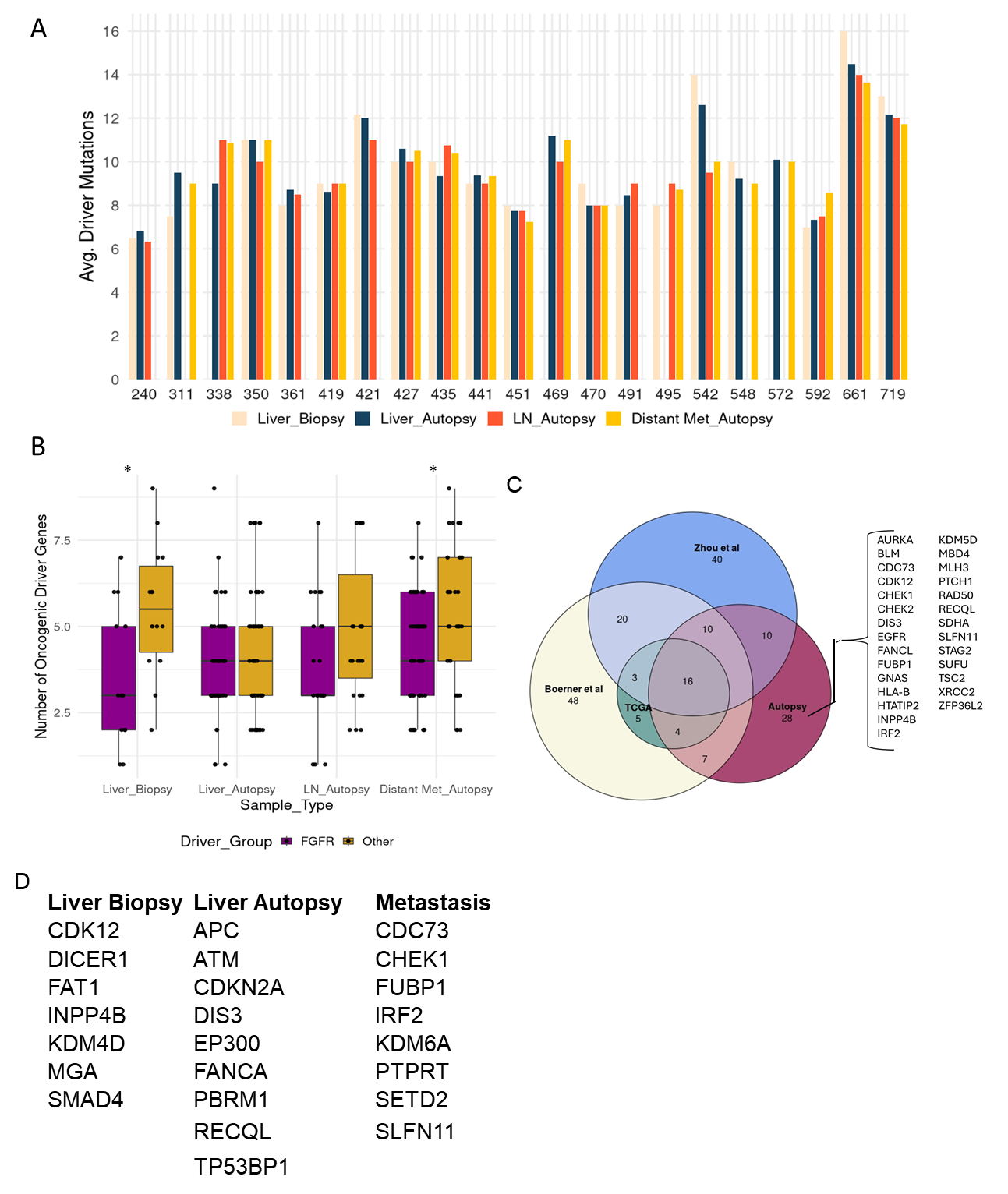

**Supplementary Figure 3. Characterization of driver mutations in iCCA Rapid Autopsy Cohort** (A) Barplot showing the average number of driver mutations per patient across sample types, including liver biopsies, liver autopsy tissue, lymph node autopsy samples, and distant metastases in each individual patient. (B) Boxplot comparing the number of oncogenic driver alterations in FGFR2-altered versus FGFR2-wild-type tumors across sample types. (Student T-test, *p < 0.05). (C) Venn diagram comparing the driver gene set identified in this autopsy cohort with published datasets (TCGA, Zhou et al., Boerner et al.). (D) Unique drivers identified that were specific to Liver Biopsy, Liver Autopsy and Distant Metastasis of all patients.

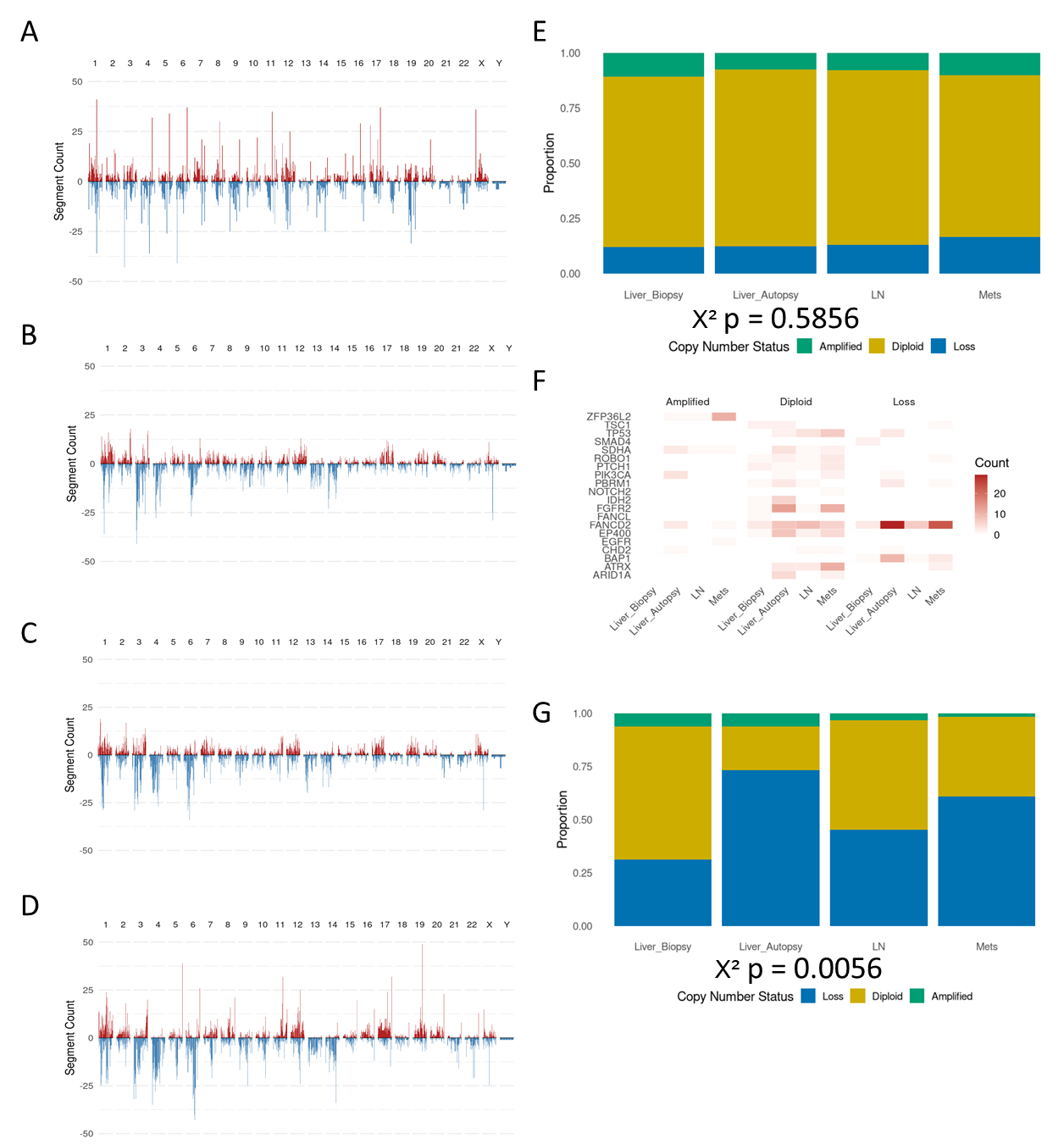

**Supplementary Figure 4. Copy Number Landscape Across Primary and Metastatic Lesions.** Genome wide copy number profiles from (A) Liver biopsy, (B) Liver Autopsy, (C) Lymph Node and (D) Metastases samples showing segment level amplifications (red) and deletions (blue) across chromosomes. (E) Relative proportions of copy number amplifications (green), diploid (yellow) and loss (blue) across sample types. (F) Frequency of copy number alterations of oncogenic drivers found in autopsy cohort across sample types. (G) Relative proportions of copy number alterations in the FANCD2 locus across sample types. There is a statistically significant difference in copy number status distribution between sites (Chi- square test, p = 0.0056).

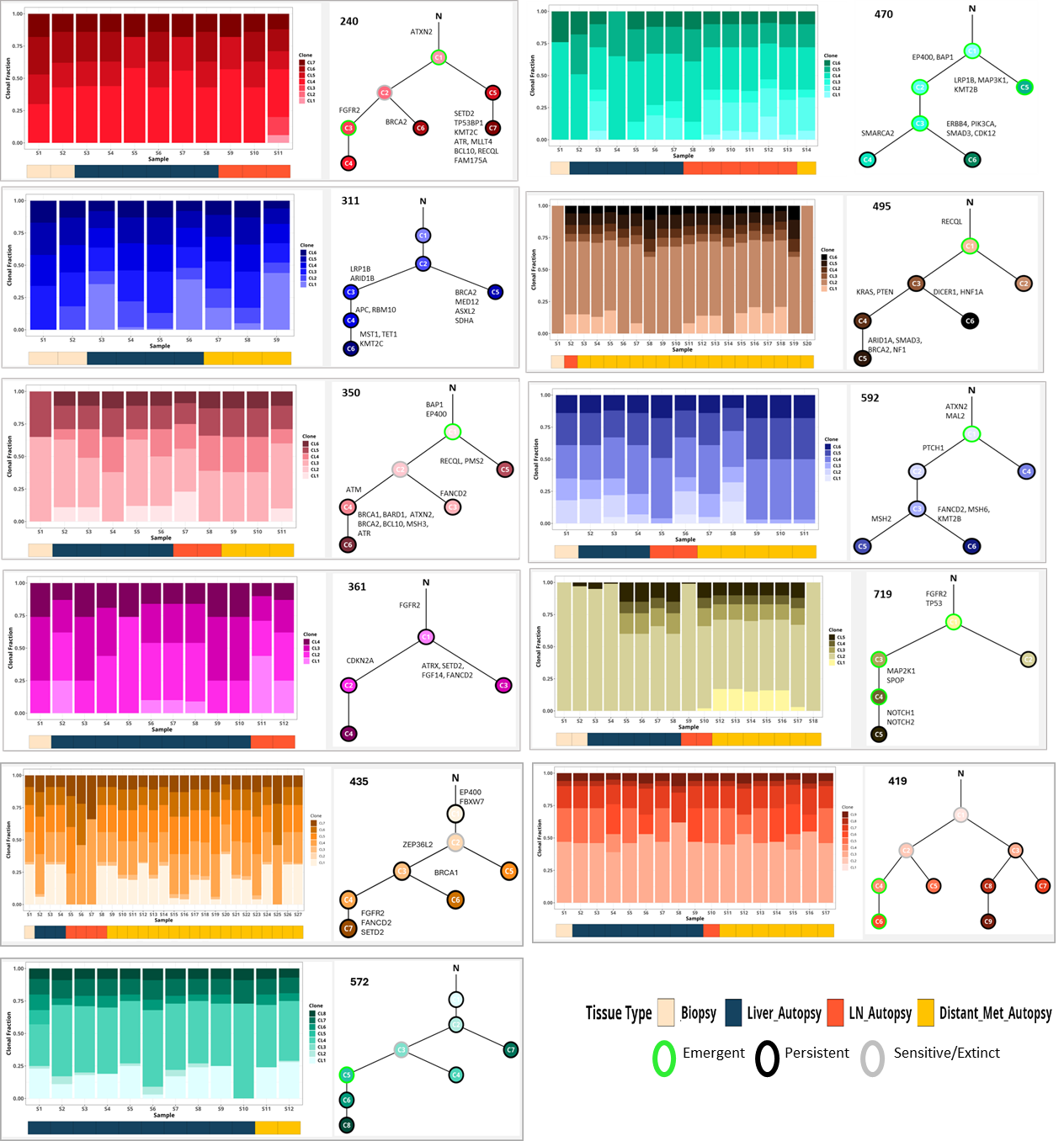

**Supplementary Figure 5.** Patient-level clonal architecture and evolutionary trajectories across multi-region samples in FGFR aberrant patients. Cancer Cell Fraction (CCF) and representative phylogeny trees as determined by Canopy for FGFR aberrant patients with major mutations annotated at branch points. For each indicated patient, the left panel shows stacked bar plots of the inferred clonal composition across all available samples as determined by Canopy (rows of bars = samples from biopsy, liver autopsy, lymph node, and distant metastases; sample class is shown by the color strip beneath each bar).

The right panel shows the corresponding phylogenetic tree inferred from multi-region WES (ASCETIC), with nodes representing clones and annotated by oncogenic events assigned to that branch (along with annotating emergent, persistent or sensitive/extinct clones).

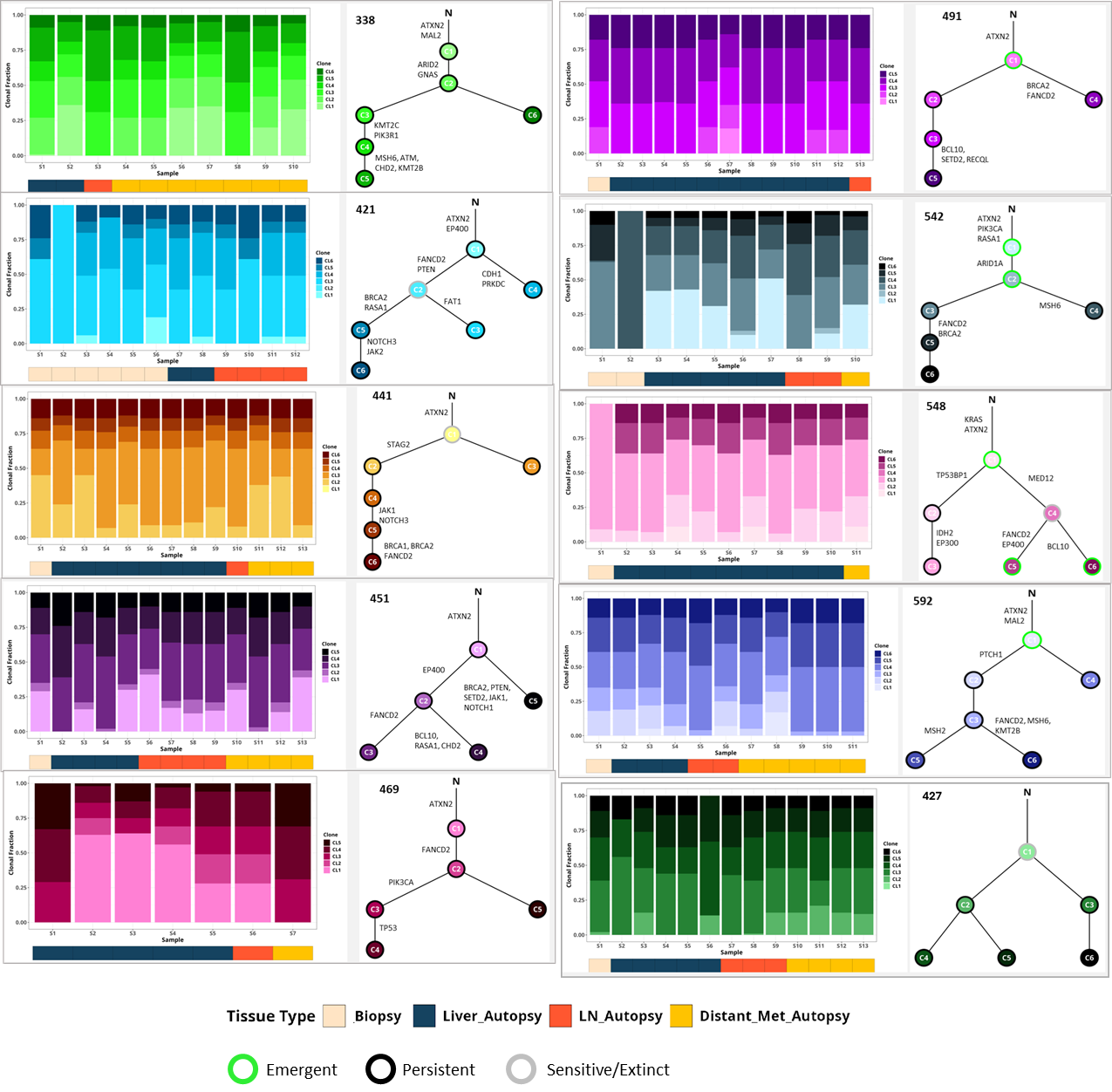

**Supplementary Figure 6.** Patient-level clonal architecture and evolutionary trajectories across multi-region samples in FGFR-WT patients. Cancer Cell Fraction (CCF) and representative phylogeny trees as determined by Canopy for FGFR aberrant patients with major mutations annotated at branch points. For each indicated patient, the left panel shows stacked bar plots of the inferred clonal composition across all available samples as determined by Canopy (rows of bars = samples from biopsy, liver autopsy, lymph node, and distant metastases; sample class is shown by the color strip beneath each bar).

The right panel shows the corresponding phylogenetic tree inferred from multi-region WES (ASCETIC), with nodes representing clones and annotated by oncogenic events assigned to that branch (along with annotating emergent, persistent or sensitive/extinct clones).

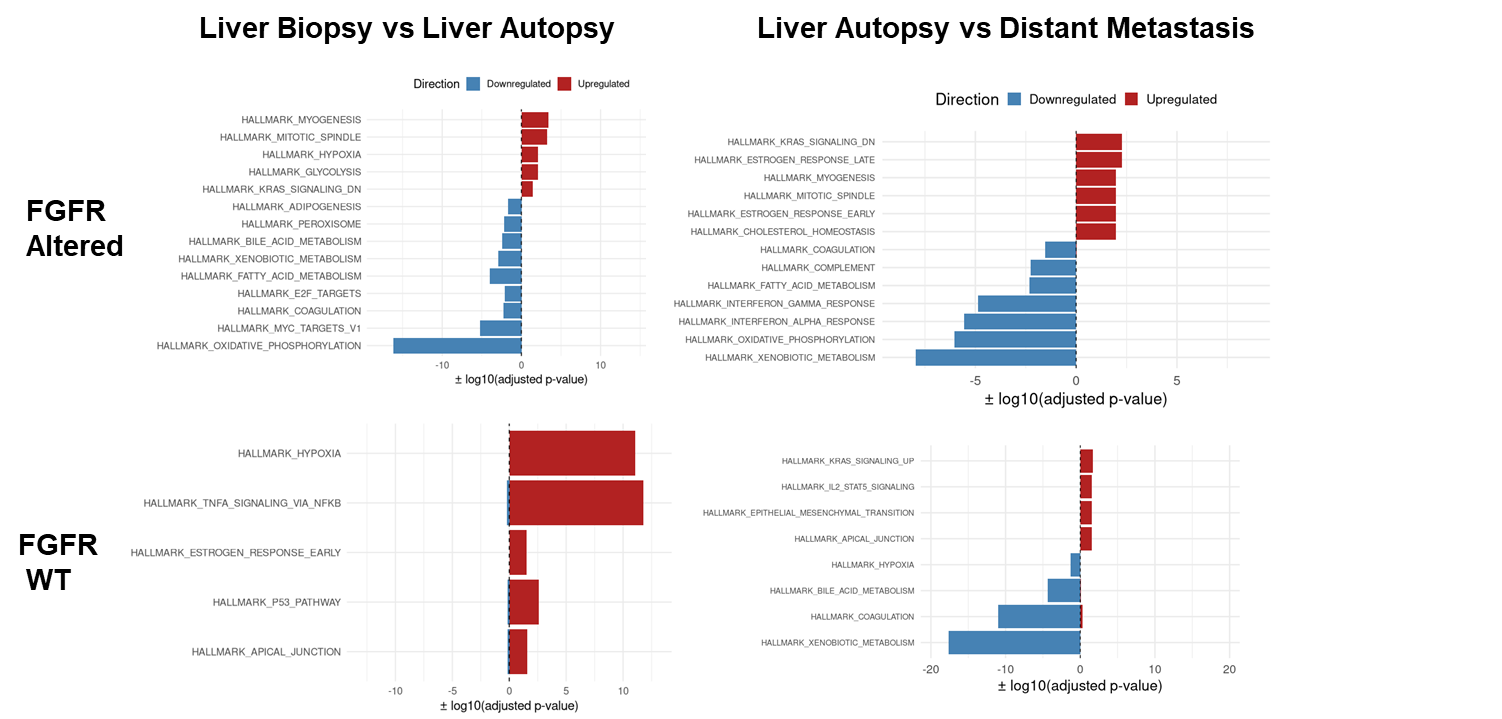

**Supplementary Figure 7. Pathway-level transcriptomic differences across sites and FGFR2 status.** Gene Set Enrichment Analysis (GSEA) of bulk RNA-seq comparing liver lesions and distant metastases. Each bar plot shows hallmark gene sets significantly upregulated (red) or downregulated (blue), with the x-axis indicating the adjusted p-value (–log10 scale) for FGFR Altered and WT Liver autopsy vs liver biopsy, Distant metastases vs liver autopsy.
